# The Parenting and Family Adjustment Scales (PAFAS) questionnaire: psychometric qualities of the parenting scale in two large Brazilian birth cohorts

**DOI:** 10.1101/2024.09.06.24313039

**Authors:** Rafaela Costa Martins, Hugo S. Gomes, Andreas Bauer, Maurício Scopel Hoffmann, Christian Loret de Mola, Elisa Rachel Pisani Altafim, Marlos Rodrigues Domingues, Joseph Murray

## Abstract

**Objective:** Analyse the psychometric properties of the Portuguese version of the PAFAS (Parenting and Family Adjustment Scales) parenting scale, using data from two large Brazilian birth cohorts.

**Methods:** The original PAFAS parenting scale, which consists of 18 items (parental inconsistency 5 items, coercive parenting 5 items, positive encouragement 3 items, and parent-child relationship 5 items) was applied in two Brazilian birth cohorts in Pelotas (ages 4 [n = 4010] and 6-7 [n = 3867]) and Rio Grande (age 3 [n = 992]). Confirmatory factor analysis was conducted, and internal consistency assessed, as well as construct validity in relation to maternal depression measured on the Edinburgh Postnatal Depression Scale.

**Results:** The model with a structure of 4 subscales showed that the global scale of parenting on the PAFAS had a good fit, but certain items did not fit well on sub-scales and were removed (2 items from parental inconsistency, 1 from coercive parenting, and 1 from positive encouragement). The original form of the parent-child relationship sub-scale was maintained. Considering the total PAFAS parenting score, we found that mothers with maternal depression had a higher likelihood of more problematic parental practices than mothers without depression.

**Conclusions:** A revised 14-item PAFAS parenting scale has good psychometric properties and we encourage its use in Brazilian populations.

## Introduction

Parenting behaviour has a fundamental role in the understanding of the development of child behaviour problems and psychopathology. For example, in two meta-analyses, parental warmth was associated with a decrease in both child externalising and internalising problems, and harsh parenting predicted increases in these outcomes over time, even accounting for prior levels of child behaviour (Pinquart, 2017b, 2017a). Another recent systematic review showed that negative parenting practices (maternal spanking and corporal punishment) negatively impacted on children’s development and behaviours at different ages (Avezum et al., 2022).

Considering that almost 90% of children live in low- and middle-income countries (LMICs) (UNICEF, 2005), parenting research is particularly needed from these world regions. Furthermore, a recently published meta-analysis found that associations of parenting practices may vary by culture (Pinquart & Kauser, 2018). For example, the association between authoritarian parenting with child externalising and internalising problems was weaker in individualistic, as opposed to collectivistic, countries. The authors propose that these differences may be explained by differences in prevalence rates across these cultural orientations. However, for example, the use of corporal punishment is widespread globally, even with research showing the negative impact on child development. Furthermore, cultural generalisations, such as Western versus Eastern culture, may not be sufficient to explain differences in parenting effects (Lansford et al., 2010). Thus, it is crucial to further investigate parenting practices across a wide range of cultural and economic settings, using measures validated and adapted for their respective context.

To monitor progress in relation to these objectives, and support research on parenting and related interventions, requires reliable and valid measures that are feasible to apply in large studies (Altafim & Linhares, 2016). However, particularly in low- and middle-income countries, this may pose a challenge, where cross-cultural adaption and psychometric validation may not always be available (Mejia et al., 2012). In Brazil, where the current study was conducted, national parenting measures have been used, such as Parental Beliefs and Caring Practices Scale (E-CPPC), the Parenting Style Inventory and the Parenting Practices Inventory for Mothers of Babies (Rodrigues et al., 2022). The use of international instruments allows intercultural comparisons and involves a careful cross-cultural validation processes (Arafat et al., 2016). In 2017, a group of researchers started the process of translating and validating the international measure Parenting and Family Adjustment scales (PAFAS) for Brazil (Santana, 2018). Sanders and colleagues (2014) had developed PAFAS as a brief and easy-to-use measure – for both clinical and public health applications. In their initial psychometric study, 30 items were retained in seven subscales across two broader scales of parenting (18 items) and family adjustment (12 items). The original 18 items of the *parenting* scale consists of four subscales, including parental inconsistency (5 items, e.g., “I deal with my child’s misbehaviour the same way all the time”), coercive parenting (5 items, e.g., “I spank (smack) my child when they misbehave”), positive encouragement (3 items, e.g., “I praise my child when they behave well”), and parent-child relationship (5 items, e.g., “I enjoy spending time with my child”). This original study identified a 4-factor solution with good internal consistency and acceptable construct and predictive validity in an Australian convenience sample, including parents with children aged 2 to 12 years. The identified factor structure was confirmed in a second study reported in the same article, using data from a different Australian convenience sample. The English version of the scale was further examined in another study of Australian parents of children with a developmental disability. Mazzucchelli et al. confirmed the 4 subscales model, albeit with 16, as opposed to 18, items. In this clinical sample, the four subscales demonstrated internal consistency and convergent validity. (Mazzucchelli et al., 2018)

To date, PAFAS has been validated in four non-English languages, including Spanish, Chinese, Indonesian, and Portuguese (Brazil). Using a convenience sample of parents from poor neighbourhoods in Panama City, Mejia and colleagues (Mejia et al., 2015) provide further support for the 4 subscales model of the parenting scale, albeit again with two items removed, with good reliability and acceptable validity. The Chinese version again supported the 4 subscales structure, after removal of three items, with satisfactory reliability and validity (Guo et al., 2016). Finally, Sumargi et al. (2018) validated the Indonesian version of PAFAS; again, after the removal of three items, they found that a model of 4 subscales best fits the data, with acceptable psychometric properties. In Brazil, a confirmatory factor analysis (CFA) of the instrument was carried out in a sample of a city of Mato Grosso do Sul in the Brazil Midwest region, which resulted in 14 items for the parenting scale (Correia et al., 2024; Santana, 2018). This model with 4-factor solution for parenting was used in different intervention studies and showed sensitivity to detect meaningful changes after the participation of mothers in parenting programs (Belotti et al., 2019; Lotto, 2023).

In addition to these validation studies, PAFAS has also been used in a study conducted in Afghanistan and in another study using data from 15 different countries^1^ (Pandya, 2018; the Afghanistan field implementation team et al., 2020). However, it should be noted that although these two studies report details on its translation, no further information is provided regarding psychometric properties. Thus, it is unclear if the measure in its original format is applicable in these diverse settings, especially considering that some modifications were required in Spain, China, Indonesia, and Brazil. In sum, although previous studies (Correia et al., 2024; Guo et al., 2016; Mejia et al., 2015; Sumargi et al., 2018) provide initial support for the PAFAS parenting scale across diverse social and cultural contexts, including Brazil, further research in large representative samples is needed. Brazil has a vast geographical area and broad diversity in socioeconomic status, cultural practices, and educational backgrounds, which may influence parenting and its measurement (Petrucci et al., 2016). Therefore, psychometric studies in diverse samples are needed to establish generality, and robustness of the measurement instrument across different populations.

To address these gaps in the literature, the current study aimed to analyse the psychometric properties of the Portuguese version of the PAFAS parenting scale, using data from two large Brazilian birth cohorts. In the 2015 Pelotas Birth Cohort, we examined (i) construct validity using confirmatory factor analysis; (ii) internal consistency using Cronbach’s alpha and McDonald’s Omega; (iii) test-retest reliability using longitudinal data from two time points; and (iv) convergent validity - testing for associations with closely related constructs. Also, we re-examined the identified factor structure using data from the 2019 Rio Grande Birth Cohort. It is important to note that the present study focuses only on the parenting practices scale and did not include the PAFAS family adjustment scale.

## Method

### Study 1: The 2015 Pelotas Birth Cohort

Pelotas is a southern Brazilian city, with approximately 344,000 inhabitants. In 2015, all live births of mothers living in the urban area of Pelotas from the five maternity wards were recruited to participate in the cohort. Between January 1st and December 31, 2015, mothers were interviewed by trained personnel. In total, 4,275 mothers and their children were included, representing 98.7% of all births in the city. (Murray et al., 2024)

These children have been followed up when they were 3, 6, 12, and 24 months old. In 2018-19, these children were followed up when they completed 4 years of age (n=2010; 95.4% follow-up rate). During this follow-up, children and their caregivers were invited to the cohort research clinic. Children’s caregivers answered standardized questionnaires applied by trained interviewers to assess socioeconomic, demographic, and behavioural information, including the parenting scale of the Brazilian version of the Parenting and Family Adjustment Scales (PAFAS). Similar procedures were applied at age 6-7 years (n=3867; 92.0% follow-up rate), but at this follow-up the version of PAFAS applied was already reduced (to 14 items) due to initial analyses of age-4 data, and a previous study from Brazil suggesting the appropriateness of using a slightly reduced version of the questionnaire (Santana, 2018).

The study was approved by the Research Ethics Committee of the Federal University of Pelotas School of Physical Education (at age 4: #26746414.5.0000.531 and at age 6-7: # 51789921.1.0000.5317) and a separate approval was obtained for specific psychological measures at age 4 from the Research Ethics Committee from Medical School of the same University (#03837318.6.0000.5317). A legal guardian was informed about the objectives of the study and were asked to sign a consent form to be eligible to participate, since participants were minors.

### Study 2: The 2019 Rio Grande Birth Cohort

Rio Grande is a city located 60km south from Pelotas. In 2019, all births from the city hospitals were identified, mothers were invited, and 99.5% participated (n=2314). Mothers of newborns weighing ≥500 grams or at least 20 weeks of gestational age were eligible to respond to a standardized interview, applied by a trained interviewer.

In 2020-2022, we followed all mothers who had a singleton liveborn in 2019 and lived in the urban area of the city. Due to the occurrence of the COVID-19 pandemic, data collection occurred remotely in a project called WebCOVID-19. During 2020, two follow-ups were completed (WebCOVID-19 waves 1 and 2; follow-up rates of 54.1% and 51.1%, respectively), and between October 2021 and May 2022, we conducted a third follow-up (WebCOVID-19 wave 3; n=992 and 48.7% follow-up rate), in which we applied the PAFAS questionnaire with the 14 questions which had already been applied at age 6-7 in the Pelotas birth cohort.

We obtained approval from the ethics committee of Universidade Federal do Rio Grande (protocol #15724819.6.0000.5324). A legal guardian was informed online about the objectives of the study and were asked to click to consent, since participants were minors.

### The PAFAS questionnaire

The original PAFAS questionnaire has 18 items on its parenting scale, with four subscales: parental inconsistency (5 items), coercive parenting (5 items), positive encouragement (3 items), and parent–child relationship (5 items). The items are statements about parenting behaviours and attitudes regarding the last four weeks, with responses indicating the frequency they occurred on a Likert-type scale ranging from 0 to 3 where 0 is “not at all”, 1 “a little”, 2 “quite a lot”, and 3 “very much”. After reverse-coding some items in order to have all items in the same direction, a sum of the 18 items ranged from 0 to 54 points, with higher scores indicating more problematic parenting behaviours. We used Santana’s (2018) previously translated version of the questionnaire to Portuguese.

### Maternal depression

We examined maternal depression as a test of construct validity in relation to PAFAS parenting scores. In the 4- and 6-7-year follow-ups of the 2015 Pelotas Birth Cohort, and in WebCOVID-19 wave 3 of the 2019 Rio Grande Birth Cohort we measured maternal depression with the Edinburgh Postnatal Depression Scale (EPDS) (Cox et al., 1987). The scale indicates the frequency of depressive symptoms over the preceding seven days. It is a screening questionnaire, with 10-items each having four possible responses, ranging in value from 0 to 3 (self-administered in the Rio Grande cohort and by interviewers in the Pelotas cohort). The Brazilian version of the questionnaire showed that a cut-off score of ≥10 identified women at risk of minor depression (Santos et al., 2007).

### Statistical analysis

We evaluated the factor structure of the PAFAS parenting scale through confirmatory factor analysis, using a robust maximum likelihood estimator in AMOS. To assess the model fit we used the comparative fit index (CFI), Parsimony comparative fit index (PCFI), Bentler-Bonett non-normed fit index (NNFI), the root-mean-square error of approximation (RMSEA), and the Akaike information criterion (AIC) was used to evaluate model fit. For the model to be considered to have an acceptable fit: the CFI and NNFI values should be above 0.95, although values above 0.90 are considered adequate; PCFI values should be above 0.80, although values above 0.60 are considered adequate, RMSEA values should be below 0.05, although values below 0.08 are considered adequate (Arbuckle, 2019; Hu & Bentler, 1999). AIC provides relevant information to compare different models, in which the lowest scores represent the models with the best fit (Mohammed et al., 2015).

Internal consistency was tested using both Cronbach’s alpha (assumes equivalent factor loadings) and McDonald’s Omega (do not assume equal factor loadings). Internal consistency values above 0.70 were considered adequate (Loewenthal, 2001). Test-retest reliability was assessed using the intraclass correlation coefficient (ICC). ICC scores above 0.75 are considered good reliability, although values between 0.5 and 0.75 are considered moderate (Koo & Li, 2016). Finally, convergent validity was assessed by testing the correlation and associations between PAFAS subscales and total PAFAS score with maternal depression, since maternal depression affects parenting behaviour (Lovejoy et al., 2000). For correlation purposes, we used maternal depression as a count variable, while in associations we used it as categorical (≥10 points) to discriminate groups of depressed or not depressed mothers according to parenting behaviour. Lastly, we used Poisson regression models with robust variance to estimate the association of maternal depression and parental practices and report incidence rate ratios.

All studies applied the questionnaire using REDCap (Research Electronic Data Capture) (Harris et al., 2009), which is a secure, web-based software platform designed to support data capture for research studies. Statistical analyses were run in SPSS and AMOS software version 28.

## Results

In the Pelotas birth cohort, data on PAFAS were available for 3970 participants (94.3% of those eligible) at age 4-years, and 3858 (91.7%) at age 6-7. In the Rio Grande birth cohort, data were available for 890 individuals (43.7%).

### Confirmatory Factor Analysis (CFA)

We started the analysis for the present study by conducting a Confirmatory Factor Analysis (CFA) with the Pelotas birth cohort analyzed when children were 4 years. The original four-factor model with 18 items showed fit indexes below the considered cut-off values, suggesting an unsatisfactory model fit (see table 1). An analysis of the items factor loadings revealed that some items presented loadings below the suggested cut-off of 0.40 (Guadagnoli & Velicer, 1988). Taking this into consideration, we have removed 2 items from the inconsistency subscale (“I follow through with a consequence (e.g. take away a toy) when my child misbehaves”/“*Quando meu/minha filho/a se comporta mal, eu atribuo uma consequência planejada (por exemplo, retiro um brinquedo*)”; λ = 0.36 and “I deal with my child’s misbehaviour the same way all the time”/“*Eu lido com o mau comportamento do/da meu/minha filho/filha da mesma maneira o tempo todo*”, λ = -0.16), one item from the coercive subscale (“I argue with my child about their behaviour and attitude”/“*Eu discuto com meu/minha filho/filha sobre seu comportamento e atitude*”, λ = 0.34), and one item from the positive encouragement subscale (“I give my child a treat, reward or fun activity for behaving well”/“*Eu dou uma guloseima, uma recompensa ou uma atividade divertida quando meu filho/minha filha se comporta bem*”, λ = 0.25).

**Table 1.** CFA model fit indexes.

| Models | CFI | PCFI | NNFI | RMSEA | AIC |
| --- | --- | --- | --- | --- | --- |
| Pelotas birth cohort |  |  |  |  |  |
| 18-item (at age 4) | 0.88 | 0.67 | 0.85 | 0.062 | 2204.04 |
| 14-item (at age 4) | 0.95 | 0.64 | 0.93 | 0.052 | 932.51 |
| 14-item (at age 4)* | 0.93 | 0.63 | 0.89 | 0.062 | 1246.67 |
| 14-item (at age 6-7) | 0.95 | 0.74 | 0.94 | 0.052 | 904.93 |
| 14-item (at age 6-7)* | 0.95 | 0.74 | 0.93 | 0.056 | 1032.50 |
| Rio Grande birth cohort |  |  |  |  |  |
| 14-item (at age 3) | 0.95 | 0.65 | 0.93 | 0.049 | 321.49 |
| 14-item (at age 3)* | 0.89 | 0.60 | 0.84 | 0.075 | 520.90 |

\*Changing the item “I enjoy giving my child hugs, kisses and cuddles” from the parent-child relationship for the positive encouragement subscale

The resulting 14-item solution presented fit scores indicative of a good model fit, except for the PCFI which presented satisfactory results. Considering previous studies analyzing psychometrics that considered changing item 15 (“I enjoy giving my child hugs, kisses and cuddles”/“*Eu gosto de dar abraços, beijos e fazer carinho no/na meu/minha filho/a*”) from parent-child relationship to positive encouragement subscale, we have carried out a new CFA which resulted in a similar, but slightly poorer, fit than the original model (see Table 1). The considered comparative fit index (i.e., AIC) also provides evidence favoring the original structure, where item 15 is included in the parent-child relationship subscale.

At age 6-7, the final 14-item version of PAFAS was applied in the Pelotas birth cohort. CFA testing the fit of the factor structure shown in Figure 1 resulted in very good factor loadings to adequate model fit (see Table 1). We also tested the model with Rio Grande birth cohort data at age 3, using 14-items, and the results showed an overall decrease in model fit. Again, comparative index provided evidence of a better fit in the original factor structure.

**Figure 1.**
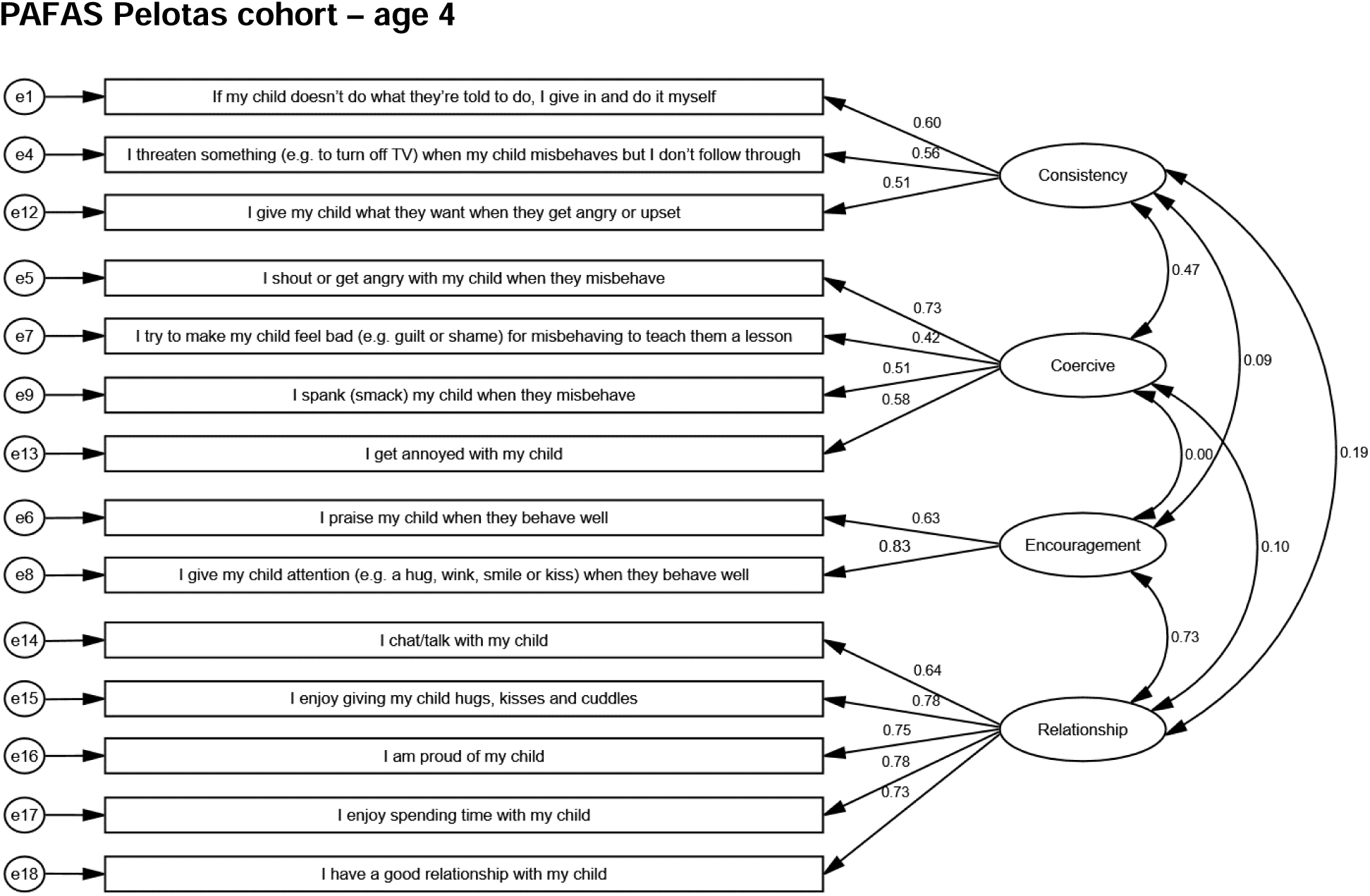
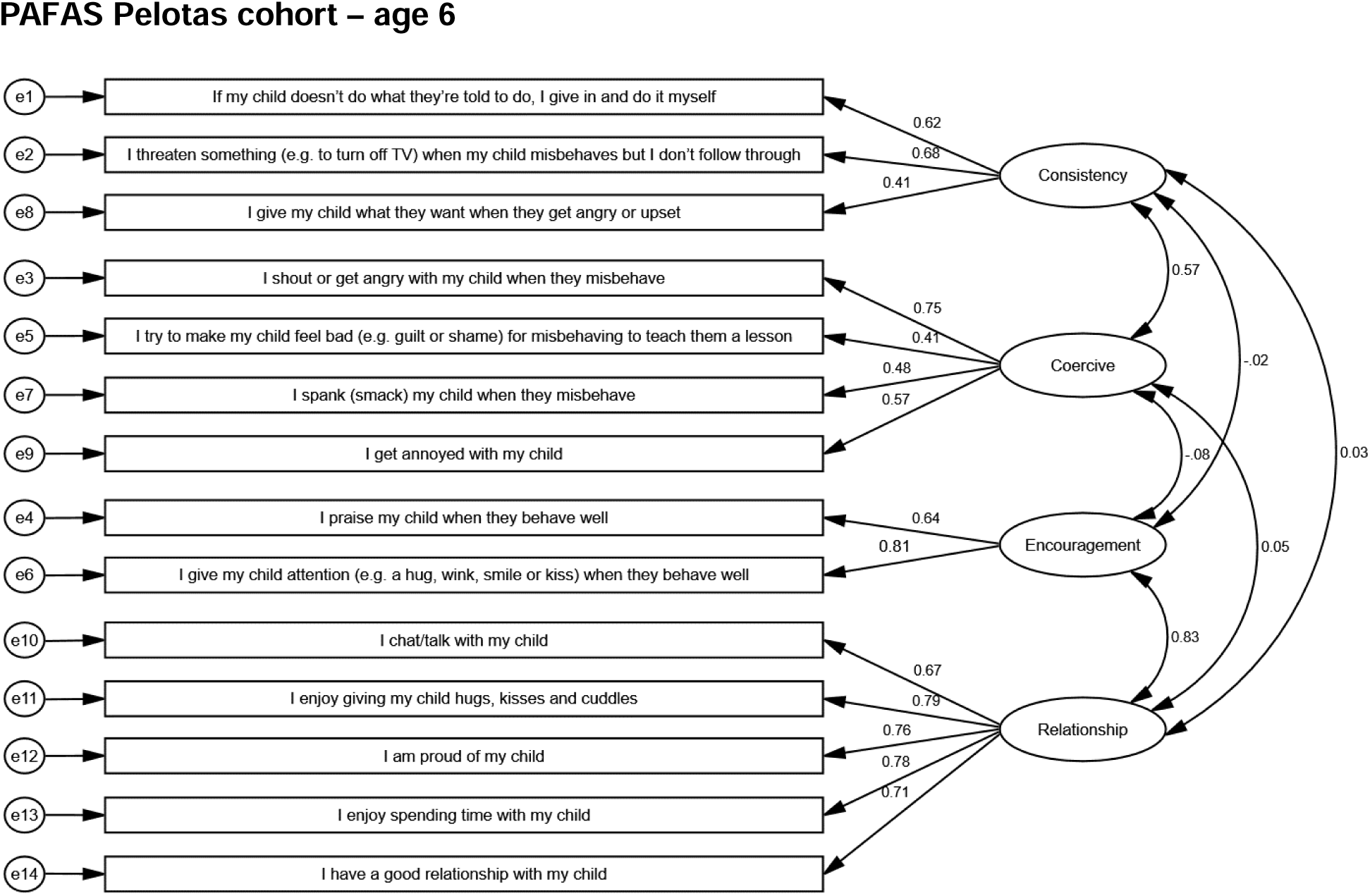
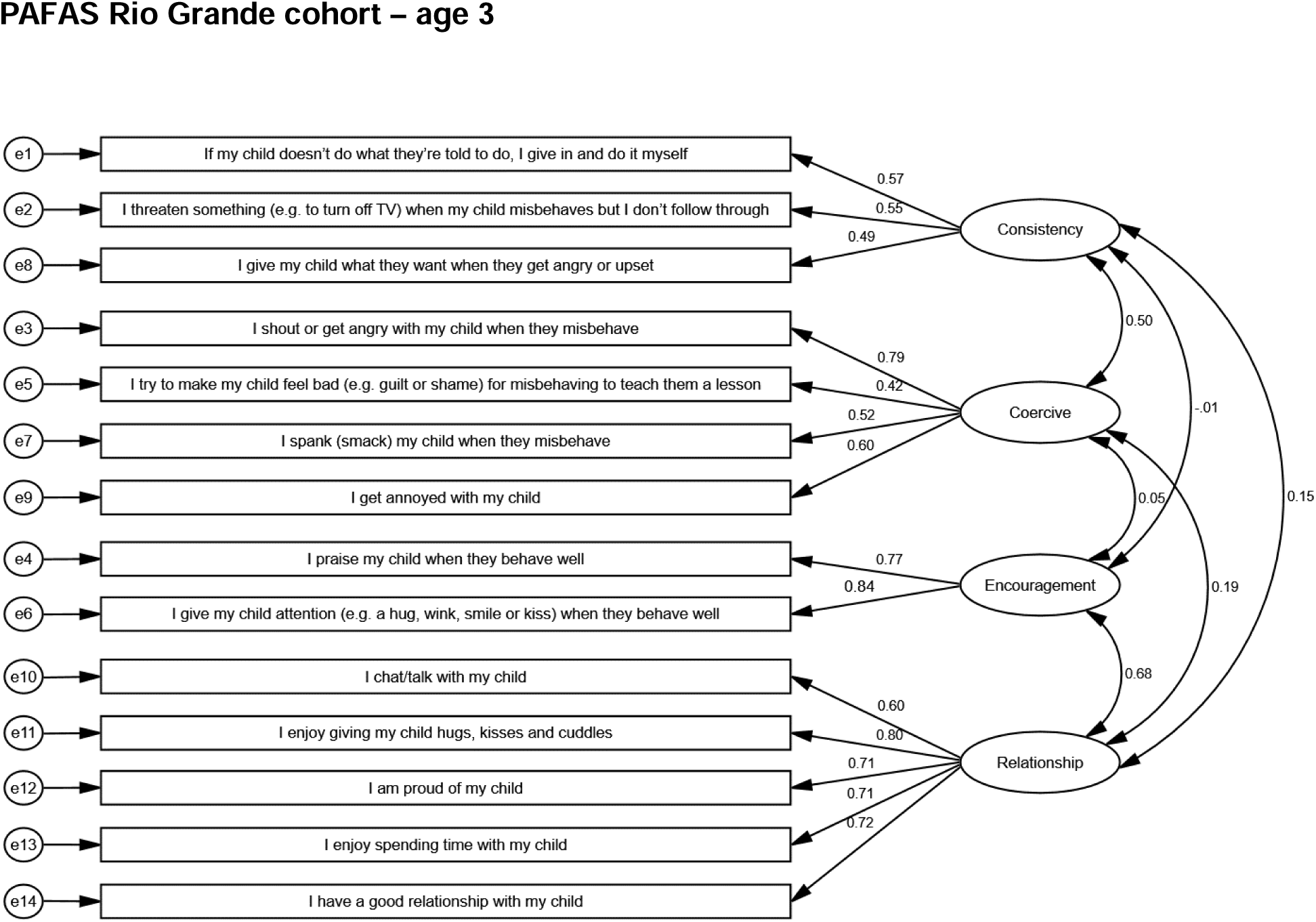
Factorial model for PAFAS items in A) Pelotas birth cohort at age 4, B) Pelotas birth cohort at age 6-7, and C) Rio Grande birth cohort.

Finally, we tested the PAFAS model fit using data from the Rio Grande birth cohort, but also switching the item “I enjoy giving my child hugs, kisses and cuddles” from the parent-child relationship for the positive encouragement subscale. Similar to the previous analysis, the original 4-factor structure with fourteen items showed very good to adequate fit scores. Again, the changes proposed in previous validation studies resulted in an overall decrease in the model fit of PAFAS in Rio Grande (see Table 1).

### Internal consistency

As illustrated in Table 2, the reliability of PAFAS was tested both using Cronbach’s alpha and omega composite reliability tests. The overall PAFAS scale presented adequate consistency (> 0.70) in both waves of Pelotas cohort, as well as in the Rio Grande cohort. Considering the internal consistency of PAFAS subscales, findings showed that the parental inconsistency subscale, as well as the coercive parenting subscale, both presented very low consistency throughout the three samples. As for the positive encouragement subscale, results for the Pelotas follow-ups at ages 4 and 6-7 presented scores slightly below the cutoff (i.e., 0.70), while ω reached the score of 0.70 at age 4. The positive encouragement subscale in the Rio Grande cohort presented acceptable internal consistency. Finally, the parent-child relationship subscale presented good reliability throughout the three samples considered.

**Table 2.** Descriptive statistics and Cronbach’s alpha for the PAFAS.

|  | Pelotas age 4<br>(n = 3,970) |  |  |  | Pelotas age 6-7<br>(n = 3,858) |  |  |  | Rio Grande age 3<br>(n = 890) |  |  |  |
| --- | --- | --- | --- | --- | --- | --- | --- | --- | --- | --- | --- | --- |
| | *M* | *SD* | $\alpha$ | $\omega$ | *M* | *SD* | $\alpha$ | $\omega$ | *M* | *SD* | $\alpha$ | $\omega$ |
| **Global scale of PAFAS** | 7.69 | 4.21 | 0.72 | 0.74 | 7.07 | 4.02 | 0.70 | 0.72 | 6.35 | 3.89 | 0.71 | 0.74 |
| **Inconsistency** | 2.00 | 1.74 | 0.57 | 0.57 | 2.23 | 1.81 | 0.58 | 0.60 | 2.40 | 1.77 | 0.55 | 0.55 |
| 1. If my child doesn't do what they're told to do, I give in and do it myself<br>*[Se meu/minha filho/a não faz o que peço, eu desisto e eu mesma faço]* | 0.86 | 0.82 |  |  | 1.02 | 0.87 |  |  | 0.99 | 0.79 |  |  |
| 2. I threaten something (e.g. to turn off TV) when my child misbehaves but I don't follow through<br>*[Quando meu/minha filho(a) se comporta mal eu ameaço (por exemplo, desligar a televisão), mas não cumpro]* | 0.82 | 0.89 |  |  | 0.95 | 0.94 |  |  | 0.80 | 0.87 |  |  |
| 8. I give my child what they want when they get angry or upset<br>*[Eu dou a meu/minha filho(a) o que ele/ela quer quando ele(a) fica com raiva ou chateado]* | 0.32 | 0.65 |  |  | 0.27 | 0.60 |  |  | 0.62 | 0.79 |  |  |
| **Coercive** | 3.59 | 2.08 | 0.65 | 0.65 | 3.09 | 1.97 | 0.64 | 0.64 | 2.69 | 1.88 | 0.67 | 0.68 |
| 3. I shout or get angry with my child when they misbehave<br>*[Eu grito ou fico braba com meu/minha filho(a) quando ele(a) se comporta muito mal]* | 1.39 | 0.86 |  |  | 1.34 | 0.86 |  |  | 1.19 | 0.79 |  |  |
| 5. I try to make my child feel bad (e.g. guilt or shame) for misbehaving to teach them a lesson<br>*[Eu tento fazer meu/minha filho(a) se sentir mal (por exemplo culpado(a) ou envergonhado(a) por se comportar mal, para lhe ensinar uma lição]* | 0.48 | 0.73 |  |  | 0.32 | 0.59 |  |  | 0.32 | 0.61 |  |  |
| 7. I spank (smack) my child when they misbehave<br>*[Eu dou um palmada no/a meu/minha filho(a) quando ele(a) se comporta mal]* | 0.78 | 0.69 |  |  | 0.58 | 0.67 |  |  | 0.40 | 0.57 |  |  |
| 9. I get annoyed with my child<br>*[Eu fico irritada com o(a) meu/minha filho(a)]* | 0.93 | 0.71 |  |  | 0.85 | 0.71 |  |  | 0.79 | 0.66 |  |  |
| **Positive encouragement** | 0.80 | 0.98 | 0.68 | 0.70 | 0.63 | 0.92 | 0.68 | 0.69 | 0.51 | 0.93 | 0.77 | 0.79 |
| 4. I praise my child when they behave well<br>*[Eu elogio meu filho filha quando ele se comporta bem]* | 0.46 | 0.59 |  |  | 0.35 | 0.54 |  |  | 0.31 | 0.56 |  |  |
| 6. I give my child attention (e.g. a hug, wink, smile or kiss) when they behave well<br>*[Eu dou atenção ao meu filho como um abraço, uma piscada de olho, um sorriso, ou um beijo quando ele/ela se comporta bem]* | 0.34 | 0.53 |  |  | 0.29 | 0.51 |  |  | 0.20 | 0.47 |  |  |
| **Parent-child relationship** | 1.31 | 1.86 | 0.85 | 0.86 | 1.11 | 1.78 | 0.85 | 0.86 | 0.75 | 1.52 | 0.81 | 0.84 |
| 10. I chat/talk with my child<br>*[Eu converso com o meu filho]* | 0.42 | 0.55 |  |  | 0.34 | 0.53 |  |  | 0.27 | 0.53 |  |  |
| 11. I enjoy giving my child hugs, kisses and cuddles<br>*[Eu gosto de dar abraços, beijos e fazer carinho no meu filho]* | 0.23 | 0.45 |  |  | 0.24 | 0.49 |  |  | 0.13 | 0.37 |  |  |
| 12. I am proud of my child<br>*[Eu sou orgulhosa do meu filho]* | 0.19 | 0.43 |  |  | 0.15 | 0.38 |  |  | 0.10 | 0.37 |  |  |
| 13. I enjoy spending time with my child<br>*[Eu gosto de passar o tempo com o meu filho]* | 0.23 | 0.45 |  |  | 0.19 | 0.42 |  |  | 0.15 | 0.38 |  |  |
| 14. I have a good relationship with my child<br>*[Eu tenho um bom relacionamento com o meu filho]* | 0.24 | 0.46 |  |  | 0.19 | 0.42 |  |  | 0.10 | 0.33 |  |  |

Note. Items 4, 6, 10, 11, 12, 13, and 14 were reversed coded.

### Construct validity

Table 3 illustrates the correlation results between the PAFAS subscales. Overall, findings show that the parental inconsistency subscale presented small to medium correlations with coercive parenting (*r*_(Pelotas age 4)_ = 0.29, *p* <0.001; *r*_(Pelotas age 6-7)_ = 0.36, *p* <0.001; *r*_(Rio Grande age 3)_ = 0.30, *p* <0.001;), while positive encouragement was largely correlated with parent-child relationship (*r*_(Pelotas age 4)_ = 0.58, *p* <0.001; *r*_(Pelotas age 6-7)_ = 0.65, *p* <0.001; *r*_(Rio Grande age 3)_ = 0.57, *p* <0.001;). Also, the parental inconsistency subscale presented small correlations with coercive parenting in both the Pelotas cohort at age 4 (*r* = 0.14, *p* <0.001) and Rio Grande at age 3 (*r* = 0.12, *p* <0.001), but no correlation was found in the Pelotas cohort at age 6-7. Other statistically significant inter-factor correlations are presented in Table 3, but with small effect sizes and inconsistencies throughout the different cohort studies.

**Table 3.** Correlations between PAFAS subscales.

|  | Parental<br>Inconsistency | Coercive<br>parenting | Positive<br>encouragement | Parent-child<br>relationship |
| --- | --- | --- | --- | --- |
| **Pelotas** (age 4 – in grey; age 6-7 – in blue) |  |  |  |  |
| Parental inconsistency | - | 0.29* | 0.05* | 0.14* |
| Coercive parenting | 0.36* | - | 0.00 | 0.09* |
| Positive encouragement | -0.02 | -0.07* | - | 0.58* |
| Parent-child relationship | 0.03 | 0.06* | 0.65* | - |
| **Rio Grande** (age 3) |  |  |  |  |
| Parental inconsistency | - | 0.30* | -0.01 | 0.12* |
| Coercive parenting |  | - | 0.05 | 0.16* |
| Positive encouragement |  |  | - | 0.57* |
| Parent-child relationship |  |  |  | - |

Note: \* p<0.05

In order to test the construct validity of the PAFAS, we carried out correlational analyses with maternal depression scores. Results illustrated in Table 4 show somewhat consistent findings throughout the different samples. Parental inconsistency (r ranging from 0.23 to 0.33) and coercive parenting (r ranging from 0.16 to 0.32) showed small to moderate positive associations with maternal depression. While no association with maternal depression was found between positive encouragement and parent-child relationship in the Rio Grande cohort, in the Pelotas cohort both at ages 4 and 6-7 results showed weak positive associations. Lastly, we carried out association analyses between maternal depression and parental practices (Table 5). Considering the four subscales of PAFAS, maternal depression measured both in Pelotas and Rio Grande cohorts was positively associated with all of them, except positive encouragement and parent-child relationship in Rio Grande birth cohort (non-significant results). In addition, considering the total PAFAS parenting score, we also found that mothers with maternal depression had a higher likelihood of more problematic parental practices (IRR = 1.37, IRR = 1.44, IRR = 1.21 for Pelotas at ages 4 and 6-7 and Rio Grande birth cohorts, respectively).

**Table 4.** Correlations of PAFAS parenting scores and maternal depression scores.

|  | Parental<br>Inconsistency | Coercive<br>parenting | Positive<br>encouragement | Parent-child<br>relationship | PAFAS<br>total |
| --- | --- | --- | --- | --- | --- |
| Pelotas cohort 4 years | 0.28* | 0.25* | 0.09* | 0.16* | 0.33* |
| Pelotas cohort 6-7 years | 0.33* | 0.32* | 0.07* | 0.14* | 0.38* |
| Rio Grande cohort | 0.23* | 0.16* | -0.02 | 0.06 | 0.20* |

Note: \* p<0.05

**Table 5.** Crude associations between PAFAS subscales and total PAFAS score with maternal depression, expressed as incidence rate ratio and their confidence intervals.

|  | Parental<br>Inconsistency | Coercive<br>parenting | Positive<br>encouragement | Parent-child<br>relationship | PAFAS total |
| --- | --- | --- | --- | --- | --- |
| Pelotas cohort 4<br>years | **1.53 (1.45 – 1.62)** | **1.28 (1.23 – 1.33)** | **1.23 (1.13 – 1.33)** | **1.47 (1.35 – 1.61)** | **1.37 (1.32 – 1.42)** |
| Pelotas cohort 6-7<br>years | **1.56 (1.49 – 1.65)** | **1.36 (1.31 – 1.42)** | **1.23 (1.11 – 1.37)** | **1.55 (1.39 – 1.73)** | **1.44 (1.39 – 1.49)** |
| Rio Grande cohort | **1.27 (1.16 – 1.40)** | **1.20 (1.09 – 1.31)** | 0.97 (0.76 – 1.24) | 1.27 (0.97 – 1.66) | **1.21 (1.12 – 1.31)** |

## Discussion

PAFAS is an international instrument for measuring parenting and family adjustment, that has demonstrated usability across different Brazilian regions (in the Southern region in the present study, and in the Midwest region, in a previous study; Correia et al., 2024). Furthermore, PAFAS has been employed in intervention studies in Southeast Brazil, highlighting its sensitivity to detect changes in coercive practices (Belotti et al., 2019; Linhares et al., 2022; Lotto et al., 2022), parental inconsistency, and positive encouragement (Belotti et al., 2019). In the current study of two population-based birth cohorts, after removing items with low factor loadings (explaining variance below 16%), overall fit indices improved, resulting in a four-factor model, with 14 items as the best solution for the PAFAS parenting scale.

We tested different PAFAS factor structures, based on previous literature and iterative analyses. Internal consistency indices between different structures demonstrated that α and ω were very similar - indicating that items presented similar factor loadings across structures. However, internal consistency indicators varied between different PAFAS parenting subscales. Parent-child relationship had high internal consistency in both cohorts, and at both ages 4 and 6-7, indicating sum scores on this subscale are useful as it represents a cohesive construct. However, Parental inconsistency had low α and ω indices, indicating that their sum scores capture additional information not due to the construct that this factor aims to measure. Parental coercive and positive encouragement subscales had borderline internal consistency scores, and were higher in the Rio Grande cohort at age 3 than in Pelotas at ages 4 or 6-7. Our study design cannot disentangle if this difference is due to cohort characteristics or age effects.

The items retained within the parental inconsistency and coercive practices subscales of the current study were consistent with those in the prior study (Correia et al., 2024). However, while one item (“I enjoy giving my child hugs, kisses and cuddles”) was included in the positive encouragement subscale in the previous study, this was retained in the parent-child relationship subscale in the current analysis, aligning with the original version. As a result, both (Correia et al., 2024) and the current study had the same final model structure for 14 items, except for the placement of a single item between two different subscales.

The four items that did not load well in either the current Brazilian samples or in previous research (Correia et al., 2024) might have problems because of the varying possible interpretations within Brazil. For instance, consider the item “I argue with my child about their behavior and attitude.” In Brazilian culture, the word “argue” would normally require an adjective or further explanation to ensure a consistent understanding between different caregivers. As written in the sentence, it remains unclear whether the practice has a positive or negative connotation, leading to potentially diverse interpretations by caregivers. Another situation arises regarding the item “I follow through with a consequence (e.g., take away a toy) when my child misbehaves”. The way this consequence is executed—whether calmly or angrily—significantly influences whether it is perceived as a positive or negative practice in any culture. Another item that demonstrated poor factor loading in the Brazilian version was the statement “I deal with my child’s misbehaviour the same way all the time.” These two items, one addressing consequences and the other concerning the approach to handling a child’s misbehaviour, were also excluded from the Panamá (Mejia et al., 2014), Chinese (Guo et al., 2016), and Indonesian (Sumargi et al., 2018) versions.

A large international literature links parental depression and different parenting behaviours, and quality of parent-child interaction (Galbally & Lewis, 2017). For instance, a prior study in Pelotas city found that depressed mothers were more likely to use harsh parenting practices and less likely to use positive parenting (Murray et al., 2023). In the current study, across both cohorts and ages, maternal depression was associated with the total PAFAS score, as well as nearly all parenting sub-scores, confirming construct validity of the questionnaire.

A strength of our study is the large samples in two different, population-based birth cohorts, assessed at multiple ages. However, some limitations should be considered. Losses in follow-ups, especially in the Rio Grande cohort, could undermine the generalisability of findings. Data came entirely from maternal reports, including the depression score used for to tests construct validity. We used established cut-offs for the CFA fit indices, although some limitations of these cut-offs are warranted. First, CFI/TLI tends to be higher (and RMSEA lower) with an increasing number of dimensions, response categories, and a lower number of items, and second, the probability of correctly rejecting a misspecified model decreases with increasing sample size (Clark & Bowles, 2018; DiStefano & Morgan, 2014; Marsh et al., 2004).

In conclusion, a global scale of parenting measured on PAFAS had a good fit across two different samples, and we encourage its use. Also, it is possible to use the separate subscales, although parental inconsistency had weak psychometric properties and is not recommended to be used alone. The strong alignment of our results with a previous Brazilian validation study (Correia et al., 2024) reaffirms the validity of the instrument in different samples within the same country.

## Data Availability

All data produced in the present study are available upon reasonable request to the authors

## Acknowledgements

This article is based on data from the study “Pelotas Birth Cohort, 2015” conducted by Postgraduate Program in Epidemiology at *Universidade Federal de Pelotas*, with the collaboration of the Brazilian Public Health Association (ABRASCO). The first phases of the 2015 Pelotas (Brazil) Birth Cohort was funded by the Wellcome Trust (095582). Funding for specific follow-up visits was also received from the *Conselho Nacional de Desenvolvimento Científico e Tecnológico* (CNPq) and *Fundação de Amparo a Pesquisa do Estado do Rio Grande do Sul* (FAPERGS) and Children’s Pastorate sponsored follow-up at twenty-four months; and FAPERGS – PPSUS, the Wellcome Trust (10735_A_18_Z), and the Bernard van Leer Foundation (BRA-2018-178) for the 4 years follow-up. At the 4 years follow-up the 2015 cohort also was funded by the Department of Science and Technology (DECIT/Brazilian Ministry of Health) and Wellcome Trust (210735_A_18_Z). The 6-7 years follow-up received funding from the Department of Science and Technology of the Ministry of Health (Department of Science and Technology (DECIT/Brazilian Ministry of Health), *Instituto Todos Pela Saúde*, Celer Biotecnologia SA, FAPERGS PqG 21/2551-0002004-0 and CNPq through public notices: 407813/2021-7, 406582/2021-1 and 406582/2021-1. This research was funded in whole, or in part, by the Wellcome Trust [Grant number 210735_A_18_Z]. Dr. Mauricio Scopel Hoffmann is supported by the United States National Institutes of Health grant R01MH120482 under his post-doctoral fellowship at UFRGS and by the Wellcome Mental Health Data Prize, granted by the Wellcome Trust (award reference 226697/Z/22/Z). For the purpose of open access, the author has applied a CC BY public copyright licence to any Author Accepted Manuscript version arising from this submission.

## Footnotes

1 India, China, Japan, Singapore, Egypt, South Africa, Saudi Arabia, France, Germany, Sweden, Italy, UK, US, Canada, and Australia.

## References

1. Altafim, E. R. P., & Linhares, M. B. M. (2016). Universal violence and child maltreatment prevention programs for parents: A systematic review. Psychosocial Intervention, 25(1), 27–38. 10.1016/j.psi.2015.10.003

2. Arafat, S., Chowdhury, H., Qusar, M., & Hafez, M. (2016). Cross Cultural Adaptation and Psychometric Validation of Research Instruments: A Methodological Review. Journal of Behavioral Health, 5(3), 129. 10.5455/jbh.20160615121755

3. Arbuckle, J. (2019). IBM® SPSS® AmosTM 26 user’s guide. [Software]. Amos Development Corporation. https://www.ibm.com/support/pages/spss-amos-26-documentation

4. Avezum, M. D. M. D. M., Altafim, E. R. P., & Linhares, M. B. M. (2022). Spanking and Corporal Punishment Parenting Practices and Child Development: A Systematic Review.

5. Trauma, Violence, & Abuse, 152483802211242. 10.1177/15248380221124243

6. Belotti, F., Altafim, E. R. P., & Linhares, M. B. M. (2019). Feasibility study of a preventive parenting program with mothers of children born preterm. Children and Youth Services Review, 107, 104526. 10.1016/j.childyouth.2019.104526

7. Clark, D. A., & Bowles, R. P. (2018). Model Fit and Item Factor Analysis: Overfactoring, Underfactoring, and a Program to Guide Interpretation. Multivariate Behavioral Research, 53(4), 544–558. 10.1080/00273171.2018.1461058

8. Correia, L. L., Altafim, E. R. P., Ferreira, P. R. D. S., Gracioli, S. M. A., Santana, L. R., & Linhares, M. B. M. (2024). Parenting and Family Adjusment Scales: Brazilian cultural adaptation. Estudos de Psicologia (Campinas), 41, e220100. 10.1590/1982-0275202441e220100

9. Cox, J. L., Holden, J. M., & Sagovsky, R. (1987). Detection of Postnatal Depression: Development of the 10-item Edinburgh Postnatal Depression Scale. British Journal of Psychiatry, 150(6), 782–786. 10.1192/bjp.150.6.782

10. DiStefano, C., & Morgan, G. B. (2014). A Comparison of Diagonal Weighted Least Squares Robust Estimation Techniques for Ordinal Data. Structural Equation Modeling: A Multidisciplinary Journal, 21(3), 425–438. 10.1080/10705511.2014.915373

11. Galbally, M., & Lewis, A. J. (2017). Depression and parenting: The need for improved intervention models. Current Opinion in Psychology, 15, 61–65. 10.1016/j.copsyc.2017.02.008

12. Guadagnoli, E., & Velicer, W. F. (1988). Relation of sample size to the stability of component patterns. Psychological Bulletin, 103(2), 265–275. 10.1037/0033-2909.103.2.265

13. Guo, M., Morawska, A., & Filus, A. (2016). Validation of the Parenting and Family Adjustment Scales to Measure Parenting Skills and Family Adjustment in Chinese Parents. Measurement and Evaluation in Counseling and Development, 074817561562575. 10.1177/0748175615625754

14. Harris, P. A., Taylor, R., Thielke, R., Payne, J., Gonzalez, N., & Conde, J. G. (2009). Research electronic data capture (REDCap)—A metadata-driven methodology and workflow process for providing translational research informatics support. Journal of Biomedical Informatics, 42(2), 377–381. 10.1016/j.jbi.2008.08.010

15. Hu, L., & Bentler, P. M. (1999). Cutoff criteria for fit indexes in covariance structure analysis: Conventional criteria versus new alternatives. Structural Equation Modeling: A Multidisciplinary Journal, 6(1), 1–55. 10.1080/10705519909540118

16. Koo, T. K., & Li, M. Y. (2016). A Guideline of Selecting and Reporting Intraclass Correlation Coefficients for Reliability Research. Journal of Chiropractic Medicine, 15(2), 155–163. 10.1016/j.jcm.2016.02.012

17. Lansford, J. E., Alampay, L. P., Al-Hassan, S., Bacchini, D., Bombi, A. S., Bornstein, M. H., Chang, L., Deater-Deckard, K., Di Giunta, L., Dodge, K. A., Oburu, P., Pastorelli, C., Runyan, D. K., Skinner, A. T., Sorbring, E., Tapanya, S., Tirado, L. M. U., & Zelli, A. (2010). Corporal Punishment of Children in Nine Countries as a Function of Child Gender and Parent Gender. International Journal of Pediatrics, 2010, 1–12. 10.1155/2010/672780

18. Loewenthal, K. M. (2001). An introduction to psychological tests and scales (2. ed). Psychology Press.

19. Lotto, C. R. (2023). Práticas parentais e comportamento de crianças: Efeitos de um programa de parentalidade presencial e remoto e dos indicadores de depressão, experiências de adversidades na infância e temperamento materno [Doutorado Direto em Saúde Mental, Universidade de São Paulo]. 10.11606/T.17.2023.tde-05012024-141753

20. Lovejoy, M. C., Graczyk, P. A., O’Hare, E., & Neuman, G. (2000). Maternal depression and parenting behavior. Clinical Psychology Review, 20(5), 561–592. 10.1016/S0272-7358(98)00100-7

21. Marsh, H. W., Hau, K.-T., & Wen, Z. (2004). In Search of Golden Rules: Comment on Hypothesis-Testing Approaches to Setting Cutoff Values for Fit Indexes and Dangers in Overgeneralizing Hu and Bentler’s (1999) Findings. Structural Equation Modeling: A Multidisciplinary Journal, 11(3), 320–341. 10.1207/s15328007sem1103_2

22. Mazzucchelli, T. G., Hodges, J., Kane, R. T., Sofronoff, K., Sanders, M. R., Einfeld, S., Tonge, B., & Gray, K. M. (2018). Parenting and family adjustment scales (PAFAS): Validation of a brief parent-report measure for use with families who have a child with a developmental disability. Research in Developmental Disabilities, 72, 140–151. 10.1016/j.ridd.2017.10.011

23. Mejia, A., Calam, R., & Sanders, M. R. (2012). A Review of Parenting Programs in Developing Countries: Opportunities and Challenges for Preventing Emotional and Behavioral Difficulties in Children. Clinical Child and Family Psychology Review, 15(2), 163–175. 10.1007/s10567-012-0116-9

24. Mejia, A., Filus, A., Calam, R., Morawska, A., & Sanders, M. R. (2015). Measuring Parenting Practices and Family Functioning with Brief and Simple Instruments: Validation of the Spanish Version of the PAFAS. Child Psychiatry & Human Development, 46(3), 426–437. 10.1007/s10578-014-0483-1

25. Mohammed, E. A., Naugler, C., & Far, B. H. (2015). Emerging Business Intelligence Framework for a Clinical Laboratory Through Big Data Analytics. Em Emerging Trends in Computational Biology, Bioinformatics, and Systems Biology (p. 577–602). Elsevier. 10.1016/B978-0-12-802508-6.00032-6

26. Murray, J., Bauer, A., Loret De Mola, C., Martins, R. C., Blumenberg, C., Esposti, M. D., Stein, A., Barros, F. C., Hallal, P. C., Silveira, M. F., Bertoldi, A. D., & Domingues, M. R. (2023). Child and Maternal Mental Health Before and During the COVID-19 Pandemic: Longitudinal Social Inequalities in a Brazilian Birth Cohort. Journal of the American Academy of Child & Adolescent Psychiatry, 62(3), 344–357. 10.1016/j.jaac.2022.07.832

27. Murray, J., Leão, O. A. D. A., Flores, T. R., Demarco, F. F., Tovo-Rodrigues, L., Oliveira, I. O., Arteche, A., Blumenberg, C., Bertoldi, A. D., Domingues, M. R., Silveira, M. F., & Hallal, P. C. (2024). Cohort Profile Update: 2015 Pelotas (Brazil) Birth Cohort Study, follow-ups from 2 to 6–7years, with COVID-19 impact assessment. International Journal of Epidemiology, 53(3), dyae048. 10.1093/ije/dyae048

28. Pandya, S. P. (2018). Spirituality to build resilience in primary caregiver parents of children with autism spectrum disorders: A cross-country experiment. International Journal of Developmental Disabilities, 64(1), 53–64. 10.1080/20473869.2016.1222722

29. Petrucci, G. W., Borsa, J. C., & Koller, S. H. (2016). Family and School in the Socioemotional Development in Childhood. Temas em Psicologia, 24(2), 403–413. 10.9788/TP2016.2-01En

30. Pinquart, M. (2017a). Associations of parenting dimensions and styles with externalizing problems of children and adolescents: An updated meta-analysis. Developmental Psychology, 53(5), 873–932. 10.1037/dev0000295

31. Pinquart, M. (2017b). Associations of Parenting Dimensions and Styles with Internalizing Symptoms in Children and Adolescents: A Meta-Analysis. Marriage & Family Review, 53(7), 613–640. 10.1080/01494929.2016.1247761

32. Pinquart, M., & Kauser, R. (2018). Do the associations of parenting styles with behavior problems and academic achievement vary by culture? Results from a meta-analysis. Cultural Diversity and Ethnic Minority Psychology, 24(1), 75–100. 10.1037/cdp0000149

33. Santana, L. R. (2018). Adaptação Transcultural e Validação da Parenting and Family Adjustment Scales (PAFAS). Universidade Federal da Grande Dourados.

34. Santos, I. S., Matijasevich, A., Tavares, B. F., Barros, A. J. D., Botelho, I. P., Lapolli, C., Magalhães, P. V. da S., Barbosa, A. P. P. N., & Barros, F. C. (2007). Validation of the Edinburgh Postnatal Depression Scale (EPDS) in a sample of mothers from the 2004 Pelotas Birth Cohort Study. Cadernos de Saúde Pública, 23(11), 2577–2588. 10.1590/S0102-311X2007001100005

35. Sumargi, A., Filus, A., Morawska, A., & Sofronoff, K. (2018). The Parenting and Family Adjustment Scales (PAFAS): An Indonesian Validation Study. Journal of Child and Family Studies, 27(3), 756–770. 10.1007/s10826-017-0926-y

36. the Afghanistan field implementation team, Haar, K., El-Khani, A., Molgaard, V., & Maalouf, W. (2020). Strong families: A new family skills training programme for challenged and humanitarian settings: a single-arm intervention tested in Afghanistan. BMC Public Health, 20(1), 634. 10.1186/s12889-020-08701-w

37. UNICEF. (2005). The state of the world’s children 2005: Childhood under threat. https://www.unicef.org/reports/state-worlds-children-2005

