## Supplementary Table 1 for "The Parenting and Family Adjustment Scales (PAFAS) questionnaire: psychometric qualities of the parenting scale in two large Brazilian birth cohorts"

Supplementary table 1. Parenting and Family Adjustment Scales (PAFAS) original (18 items) and reduced (14 items) questionnaire in English and Portuguese

**Parenting and Family Adjustment Scale (PAFAS)**

***[Escala De Parentalidade e Ajustamento Familiar]***

Please read each statement and select a number 0, 1, 2 or 3 that indicates how true the statement was of you over the past four weeks. There are no right or wrong answers. Do not spend too much time on any statement.

*[Por favor, leia cada afirmação e assinale com um círculo o número da escala que mostra até que ponto cada afirmação é verdadeira, considerando as últimas quatro semanas. Não há respostas certas ou erradas. Não gaste muito tempo com cada afirmação.]*

|  | How true is this for you?  *[Até que ponto cada afirmação é verdadeira para você?]* | | | |
| --- | --- | --- | --- | --- |
|  | Not at all  *[Nem um pouco]* | A little  *[Um pouco (algum tempo)]* | Quite a lot *[Bastante (boa parte do tempo)]* | Very much *[Muito (a maior parte do tempo)]* |
| 1. If my child doesn’t do what they’re told to do, I give in and do it myself *[Se meu/minha filho/a não faz o que peço, eu desisto e eu mesma faço]* | 0 | 1 | 2 | 3 |
| 2. I give my child a treat, reward or fun activity for behaving well *[Eu dou uma guloseima, uma recompensa ou uma atividade divertida quando meu filho se comporta bem]* | 0 | 1 | 2 | 3 |
| 3. I follow through with a consequence (e.g. take away a toy) when my child misbehaves *[Quando meu filho se comporta mal, eu atribuo uma consequência planejada (por exemplo, retiro um brinquedo)]* | 0 | 1 | 2 | 3 |
| 4. I threaten something (e.g. to turn off TV) when my child misbehaves but I don’t follow through *[Quando meu/minha filho(a) se comporta mal eu ameaço (por exemplo desligar a televisão), mas não cumpro]* | 0 | 1 | 2 | 3 |
| 5. I shout or get angry with my child when they misbehave *[Eu grito ou fico braba com meu/minha filho(a) quando ele(a) se comporta muito mal]* | 0 | 1 | 2 | 3 |
| 6. I praise my child when they behave well *[Eu elogio meu/minha filho(a) quando ele/ela se comporta bem]* | 0 | 1 | 2 | 3 |
| 7. I try to make my child feel bad (e.g. guilt or shame) for misbehaving to teach them a lesson *[Eu tento fazer meu/minha filho(a) se sentir mal (por exemplo culpado/a ou envergonhado/a) por se comportar mal, para lhe ensinar uma lição]* | 0 | 1 | 2 | 3 |
| 8. I give my child attention (e.g. a hug, wink, smile or kiss) when they behave well *[Eu dou atenção a meu/minha filho(a) como um abraço, uma piscada de olho, um sorriso, ou um beijo quando ele/ela se comporta bem]* | 0 | 1 | 2 | 3 |
| 9. I spank (smack) my child when they misbehave *[Eu dou um palmada no/a meu/minha filho(a) quando ele(a) se comporta mal]* | 0 | 1 | 2 | 3 |
| 10. I argue with my child about their behaviour/attitude *[Eu discuto com meu filho sobre seu comportamento/attitude]* | 0 | 1 | 2 | 3 |
| 11. I deal with my child’s misbehaviour the same way all the time *[Eu lido com o mau comportamento do meu filho da mesma maneira, o tempo todo]* | 0 | 1 | 2 | 3 |
| 12. I give my child what they want when they get angry or upset *[Eu dou a meu/minha filho(a) o que ele/ela quer quando ele(a) fica com raiva ou chateado/a]* | 0 | 1 | 2 | 3 |
| 13. I get annoyed with my child *[Eu fico irritada com o(a) meu/minha filho(a)]* | 0 | 1 | 2 | 3 |
| 14. I chat/talk with my child *[Eu converso com meu/minha filho(a)]* | 0 | 1 | 2 | 3 |
| 15. I enjoy giving my child hugs, kisses and cuddles *[Eu gosto de dar abraços, beijos e fazer carinho no(a) meu/minha filho(a)]* | 0 | 1 | 2 | 3 |
| 16. I am proud of my child *[Eu sou orgulhosa do/da meu/minha filho(a)]* | 0 | 1 | 2 | 3 |
| 17. I enjoy spending time with my child *[Eu gosto de passar o tempo com o/a meu/minha filho(a)]* | 0 | 1 | 2 | 3 |
| 18. I have a good relationship with my child *[Eu tenho um bom relacionamento com o/a meu/minha filho(a)]* | 0 | 1 | 2 | 3 |

Note: Translation of the instrument conducted by Correia et al., 2024, authorized by the authors of PAFAS. Grey cells were removed in the reduced version.
